# Assessment of Rational Use of Antibiotics: A Nationwide Cross-Sectional Study among People of Nepal

**DOI:** 10.1101/2022.07.11.22277488

**Authors:** Sitaram Khadka, Sulochana Khadka, Gopal Kumar Yadav, Santoshi Giri, Arun Sharma, Rinku Joshi, Kapil Amgain

**Author notes:** **Correspondence:** (GKY).

## Abstract

**Introduction:** This research was conducted with the objective to accentuate antimicrobial misuse across knowledge, behaviour and practice domains among general people of Nepal.

**Materials and Methods:** It was a nationwide cross-sectional survey conducted among 385 participants in Nepal from February 2022 to May 2022. Statistical analysis was done through SPSS® v21 and MedCalc for Windows v12.3.0. Modified Bloom’s cut-off point was utilized to categorize the participants’ overall knowledge, behaviour, and practice. The chi-square test and odds ratio (OR) using binary logistic regression at 95% CI, and Spearman’s rank correlation coefficient test (r) was calculated wherever appropriate.

**Results:** More than three-fifths of the participants (248, 64.42%) demonstrated good behaviour, whereas less than half of the participants showed good knowledge (137, 35.58%) and practice (161, 41.82%) about rational use of antibiotics. Health professionals had higher knowledge (OR: 1.07, 95% CI: 0.70-1.62) and good behaviour (OR: 0.42, 95% CI: 0.27-0.64) than other professions (P-value< 0.05). Those with higher income (≥ 50,000 NRs) had good behaviour (OR: 3.37, 95% CI: 1.65-6.87) and good practice (OR: 2.58, 95% CI: 1.47-4.50) scores than those with less monthly income (P-value< 0.05). Similarly, higher educational degrees viz., master and/or above had good behaviour (OR: 4.13, 95% CI: 2.62-6.49) and good practice scores (OR: 2.55, 95% CI: 1.68-3.87). Additionally, there were significant positive correlations between knowledge (K), behaviour (B) and practice (P) scores (r = 0.331 for K & B, r = 0.259 for K & P, and r = 0.618 for B & P respectively; P-value< 0.05).

**Conclusions:** Our findings imply the demand of effective legislature, strict enforcement of the drug act and proper implementation of plans and policies to curb the antibiotic misuse. Lack of execution of existing laws and unawareness of the public lead to extravagant use of antibiotics.

## Introduction

The irrational use of antimicrobials including antibiotics is a global issue. The use of antimicrobials and their acquisition from pharmacies without prescription is on the rise in low-and middle-income countries (LMICs) [1,2]. Inappropriate antibiotic prescription is also widely reported all over the world, including the developed countries [3]. These practices consequently promote irrational use of antimicrobials which has long-term effects on patients’ health [3]. The World Health Organization (WHO) Global Strategy for Containment of Antimicrobial Resistance outlines appropriate antimicrobial use as the cost-effective use of antimicrobials that maximizes clinical therapeutic effect whilst minimizing drug-related toxicity and development of antimicrobial resistance (AMR) [4].

Antimicrobials are the crucial for the treatment of varieties of infections which came into practice after Alexander Fleming discovered penicillin in 1928 [5]. AMR can be attributed to irrational use of antimicrobials; unnecessary, suboptimal (duration, frequency, indication, dose), and extensive use of broad-spectrum antimicrobials [6,7].AMR is one of the greatest threats to public health that is responsible for increased morbidity and mortality as well as the augmented healthcare costs [8,9].The WHO has launched a Global Action Plan (GAP) in 2015 to initiate evidence-based prescribing through effective, rapid, and low-cost diagnostic tools to optimize the use of antimicrobials [3]. Nepal has also developed National Antibiotic Resistance Containment Action Plan (NAP), 2016 on the basis of GAP. However, its implementation is still a challenge as the majority of the health care workers are unaware of this action plan and its utilization is very low [10].

Despite these efforts, the volume of antimicrobials use is ever-increasing worldwide, especially in LMICs with India in the first and China in the second position [11,12]. The incidence of AMR is more prevalent in LMICs due to poor enforcement of laws and a lack of substantial surveillance systems [13]. Nepal is no different. However, Nepal has made significant effort for the rational use of medicines (RUM), especially antimicrobials. National Antibiotics Treatment Guidelines, 2014 and Antimicrobial Resistance Containment Guideline in 2019 have been released [14]. Additionally, government and stakeholders have formed a multisectoral committee to jointly work on AMR. Nevertheless, there is no sufficient surveillance system for tracking current antibiotic use and its resistance pattern in Nepal [10]. Some national/international non-governmental organizations (NGOs/INGOs) such as Family Health International 360 (FHI 360), Nepal Public Health Foundation-Global Antibiotic Resistance Partnership (NPHF-GARP), Nepal Public Health Research and Development Center (PHRD Nepal), and Alliance for Prudent Use of Antibiotics (APUA) are working on surveillance as well as awareness and educational interventions for AMR containment in Nepal [10]. Although individuals and institutions are making sporadic attempts in all domain of AMR, there is a lack of coordinated action. Little research and published literature are not sufficient enough to elucidate the current scenario.

AMR is a serious complication in Nepal. It is really difficult to report exact trends of antibiotic use and its resistance. Therefore, this research was carried out with an objective to assess the knowledge, behaviour and practice of antibiotics misuse among general population in the context of Nepal so that effective interventions can be implemented to promote rational drug therapy.

## Materials and Methods

### Ethics approval

The research had been performed in accordance with the Declaration of Helsinki. The ethics approval was granted by the Institutional Review Committee (IRC) of the Nepalese Army Institute of Health Sciences, Kathmandu, Nepal (Reg. No 572). Informed e-consent to participate in the study was obtained from participants.

### Study design, setting and study population

A nationwide cross-sectional survey was carried out in Nepal from 14^th^February 2022 to 15^th^May 2022, principally targeting the assessment about the antibiotic use patterns in general population in accordance to the guidelines of Strengthening of the Reporting of Observational Studies in Epidemiology (STROBE) [15]. Nepal is a landlocked country in the south-east Asia situated between China and India. It has an area of 147,516 km^2^ and an estimated population of 29, 192,480 as per census 2021. The flow diagram depicting the participants inclusions has been drawn below and the final sample size was 385 (Figure 1).

**Figure 1:**
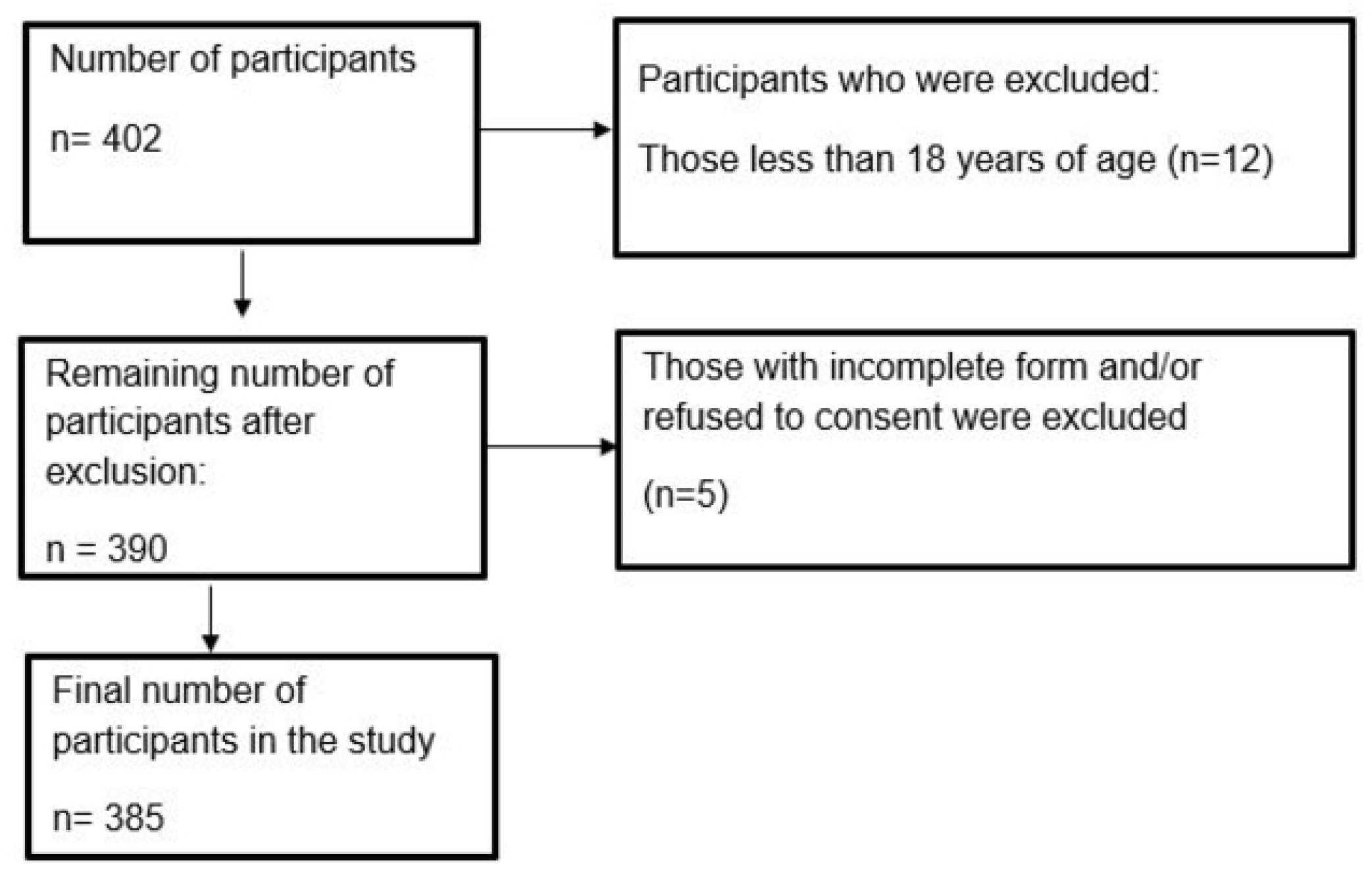
Flow diagram showing the selection of the study participants.

### Study tool and data collection

A semi-structured questionnaire was developed in English as well as in the Nepali language comprising of five sections; information and informed consent, demographics of participants, knowledge, behaviour, and practices on antibiotics use on a five-point Likert scale (strongly disagree, disagree, neutral, agree, strongly agree). The questionnaire was validated by expert review team of Nepal Army Institute of Health Sciences, Kathmandu, Nepal. It was pretested on 20 participants, which were not included in the final data set. Then, the pretested and verified questionnaire was sent to the participants using the convenience sampling method via different online medias (Messenger, Whatsapp, Viber, etc.) in the form of Google form. The first page of the “google form questionnaire” contained the “Information and informed consent sheet” to obtain e-consent after explanation of objectives of the study and voluntary nature of participation. The subsequent pages contained questions on knowledge, behaviour and practices of antibiotics use.

### Data management and statistical analysis

The response to the online survey was extracted from google docs as excel 2019 v16.0 (Microsoft, WA, USA) and exported to IBM SPSS® v21 (IBM, Armonk, NY) and MedCalc for Windows v 12.3.0 (MedCalc-Software, Mariakerke Belgium) for the further analysis.

The knowledge and behaviours item consist of five questions (maximum score 25) while the practices item consist of ten questions (maximum score 50). The statements which opposed the notion of knowledge, behaviours and/or practices were graded 5 points for strongly disagree and 1 point for strongly agree, and accordingly rest responses of disagree, neutral/unsure and agree to 4, 3 and 2 points in decremental order respectively. Similarly, the statements which supported the notion of knowledge, behaviour and/or practices were graded 1point for strongly disagree and 5 points for strongly agree, and accordingly rest responses of disagree, neutral/unsure and agree to 2, 3 and 4 points respectively.

The total scores of knowledges, behaviours, and practices were calculated and recoded into different categorical variables. The good [(≥ 80% *of* 25 = 20) for knowledge and behaviour items, and (≥ 80% *of* 50 = 40) for practice items], and the moderate to poor group (<80%) were categorized for each knowledge, behaviours and practices items based on modified Bloom’s cut off criteria [16]. Socio-demographic characteristics of participants were presented as frequency and proportions. The chi-square test was used to test for group differences. For binary logistic regression analyses, odds ratio (OR) was calculated at 95% confidence intervals (95% CI). Box plots were drawn for the distribution of knowledge, behaviour, and practice scores based on education level and areas of work. Spearman’s rank correlation coefficient test was used to assess the relationships among the knowledge, behaviour, and/or practice scores.

## Results

### Socio-demographic data

Maximum participants were of age below 40 years (360, 93.51%), and more than two-fifths were male (159, 41.30%). Most participants had educational qualifications of higher secondary and above level (365, 94.80%). More than one-fifth of the participants were from a rural area (80. 20.58%). More than three-fifths respondents were health professionals (240, 62.34%). The majority of the participants, more than 80% (323, 83.89), had income less than 50,000 NRS. Few participants were smokers (15, 3.89%). More than one-fifth had co-morbid state (63, 16.36%). (Table 1)

**Table 1:**
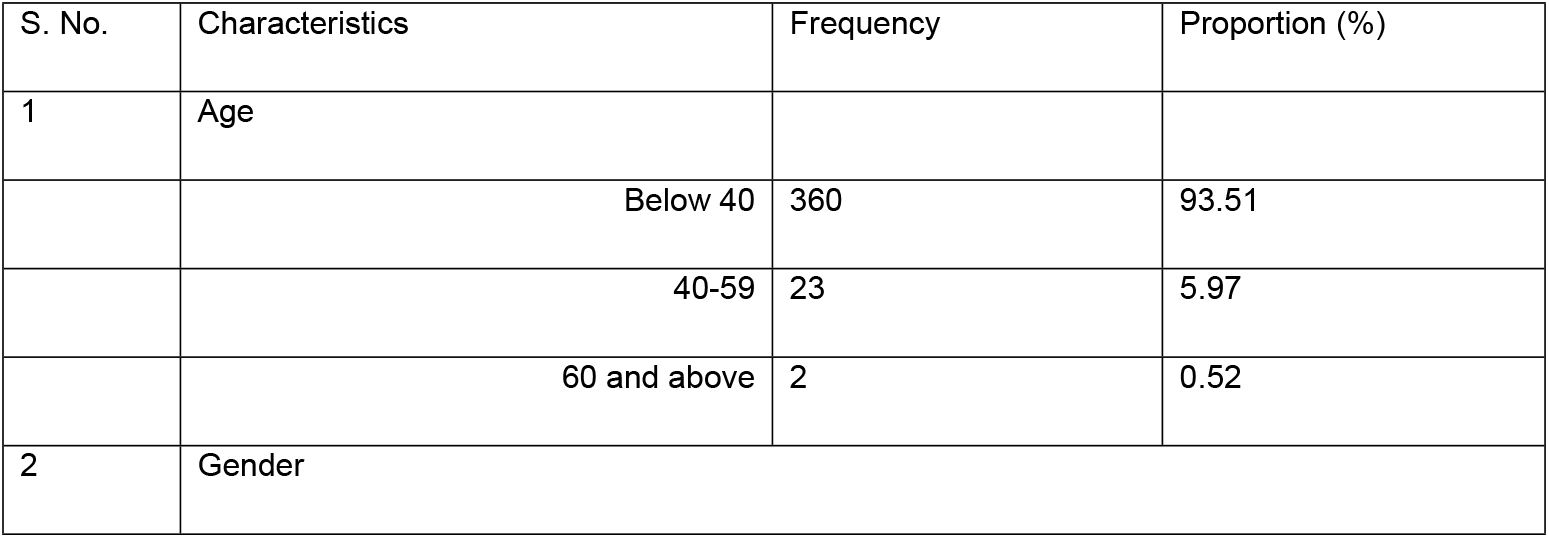
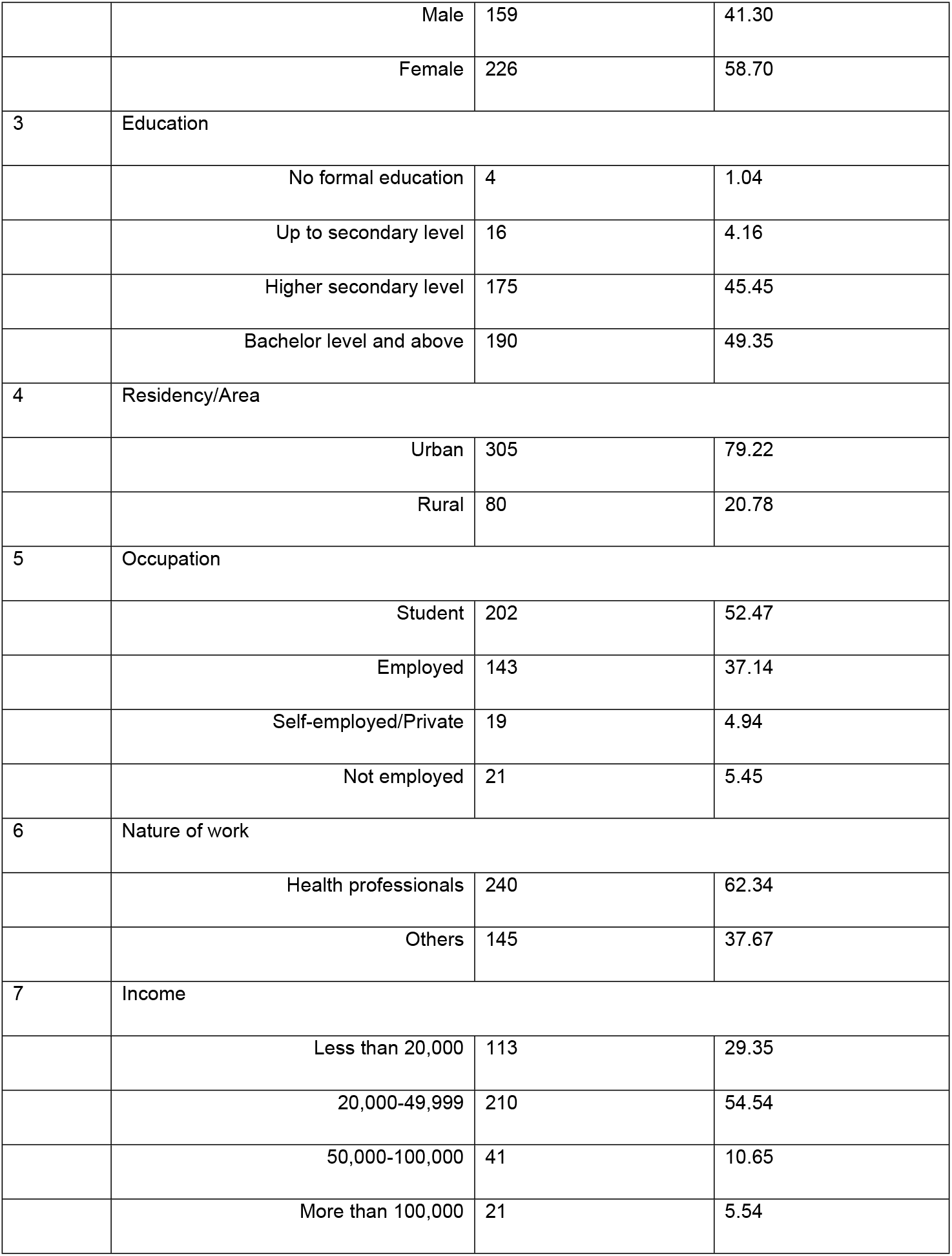
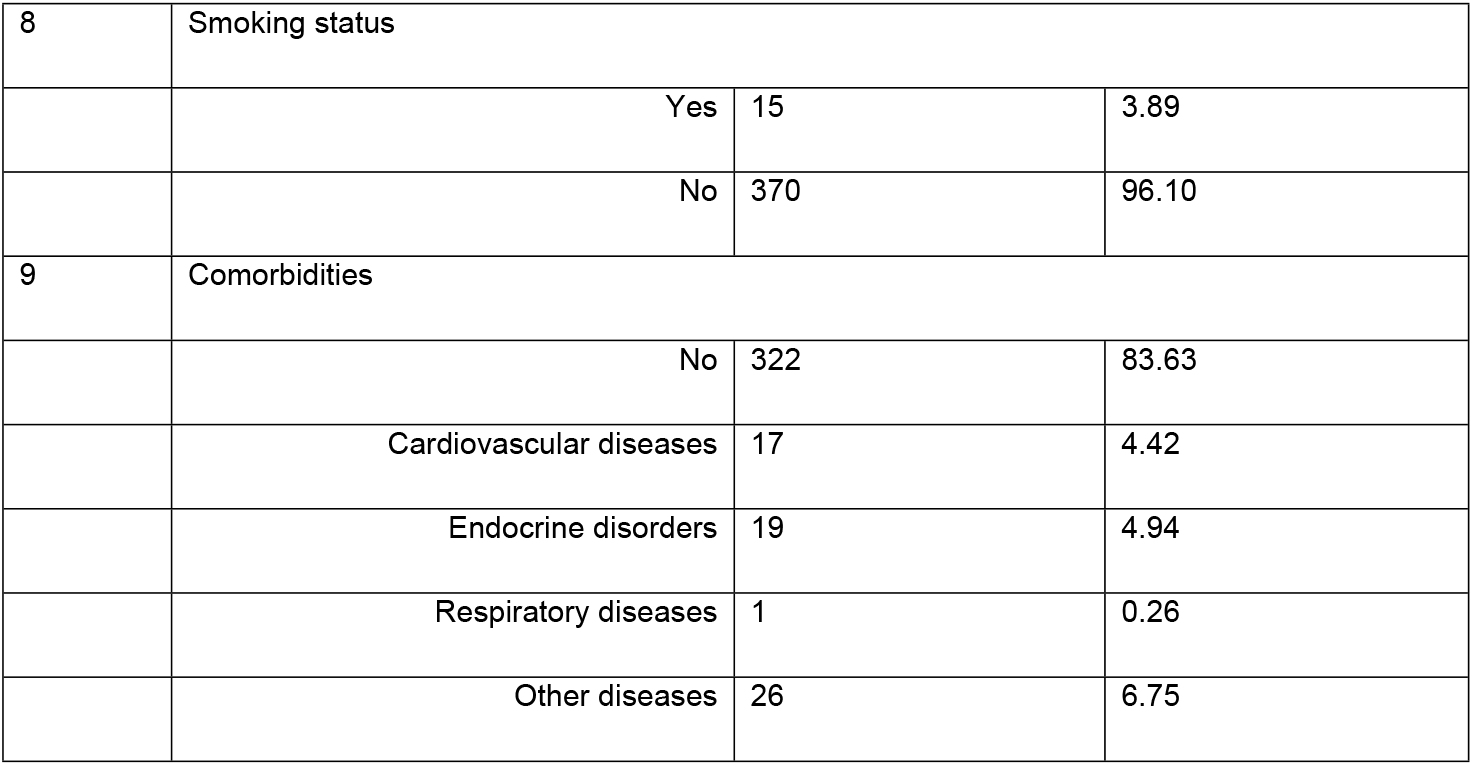
Socio-demographics characteristics of the participants (N= 385)

| S. No. | Characteristics | Frequency | Proportion (%) |
| --- | --- | --- | --- |
| 1 | Age |  |  |
|  | Below 40 | 360 | 93.51 |
|  | 40-59 | 23 | 5.97 |
|  | 60 and above | 2 | 0.52 |
| 2 | Gender |  |  |
|  | Male | 159 | 41.30 |
|  | Female | 226 | 58.70 |
| 3 | Education |  |  |
|  | No formal education | 4 | 1.04 |
|  | Up to secondary level | 16 | 4.16 |
|  | Higher secondary level | 175 | 45.45 |
|  | Bachelor level and above | 190 | 49.35 |
| 4 | Residency/Area |  |  |
|  | Urban | 305 | 79.22 |
|  | Rural | 80 | 20.78 |
| 5 | Occupation |  |  |
|  | Student | 202 | 52.47 |
|  | Employed | 143 | 37.14 |
|  | Self-employed/Private | 19 | 4.94 |
|  | Not employed | 21 | 5.45 |
| 6 | Nature of work |  |  |
|  | Health professionals | 240 | 62.34 |
|  | Others | 145 | 37.67 |
| 7 | Income |  |  |
|  | Less than 20,000 | 113 | 29.35 |
|  | 20,000-49,999 | 210 | 54.54 |
|  | 50,000-100,000 | 41 | 10.65 |
|  | More than 100,000 | 21 | 5.54 |
| 8 | Smoking status |  |  |
|  | Yes | 15 | 3.89 |
|  | No | 370 | 96.10 |
| 9 | Comorbidities |  |  |
|  | No | 322 | 83.63 |
|  | Cardiovascular diseases | 17 | 4.42 |
|  | Endocrine disorders | 19 | 4.94 |
|  | Respiratory diseases | 1 | 0.26 |
|  | Other diseases | 26 | 6.75 |

### Knowledge assessment

The responses are reported in **Supplementary Table 1**. The median knowledge score was 18 (25^th^ to 75^th^ percentiles: 17-20). More than one-third of the participants (137, 35.58%) had good knowledge regarding the RUM. The health professionals had 1.79 times higher odds of good knowledge than other professionals (OR 0.56, 95% CI 0.36-0.88) (Tables 2 and 3).

**Table 2:** Knowledge, behaviour, and practice scores of the participants.

| SN | Characteristics | Scores/Values |
| --- | --- | --- |
| 1 | Knowledge |  |
|  | Median (Min-Max) | 18 (13-25) |
|  | Q1-Q3 | 17-20 |
| | Mean ± SD | 18.51 ± 2.19 |
|  | Good knowledge | 35.58% (137/385) |
| 2 | Behaviour |  |
|  | Median (Min-Max) | 21 (8-25) |
|  | Q1-Q3 | 18-23 |
| | Mean ± SD | 20.73 ± 3.11 |
|  | Good behaviour | 64.42% (248/385) |
| 3 | Practice |  |
|  | Median (Min-Max) | 38 (12-50) |
|  | Q1-Q3 | 34-43 |
| | Mean ± SD | 37.73 ± 6.93 |
|  | Good practice | 41.82% (161/385) |

**Table 3:**
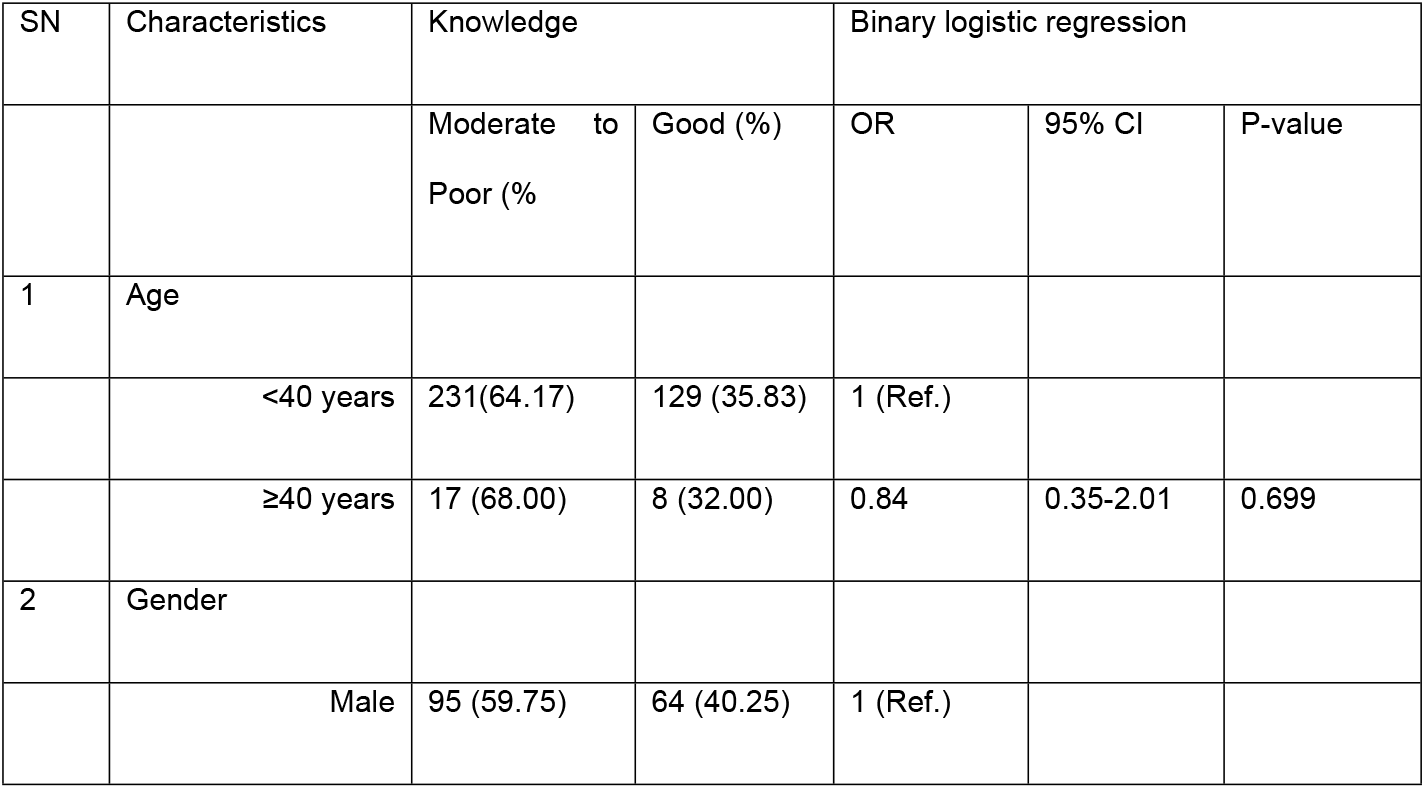
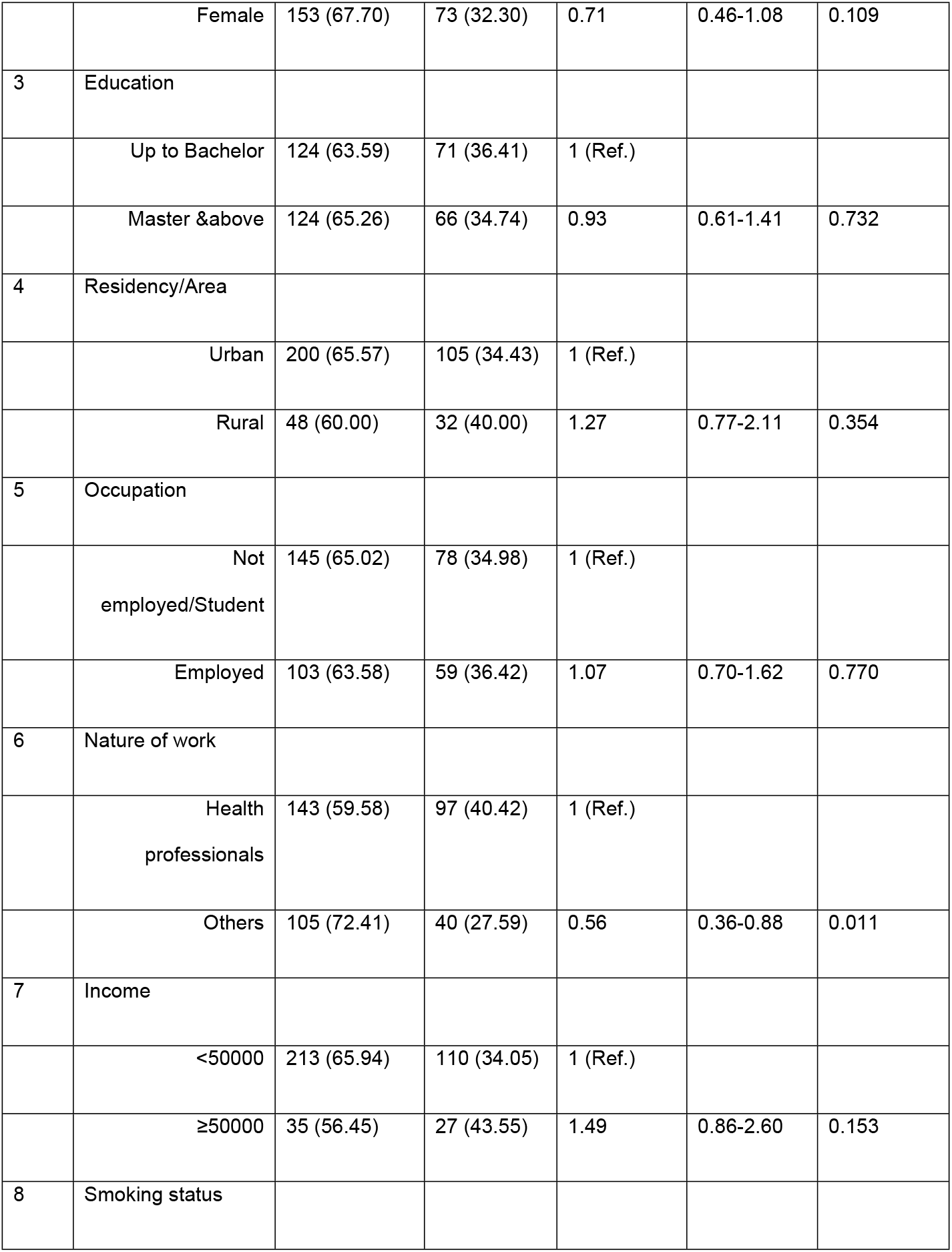
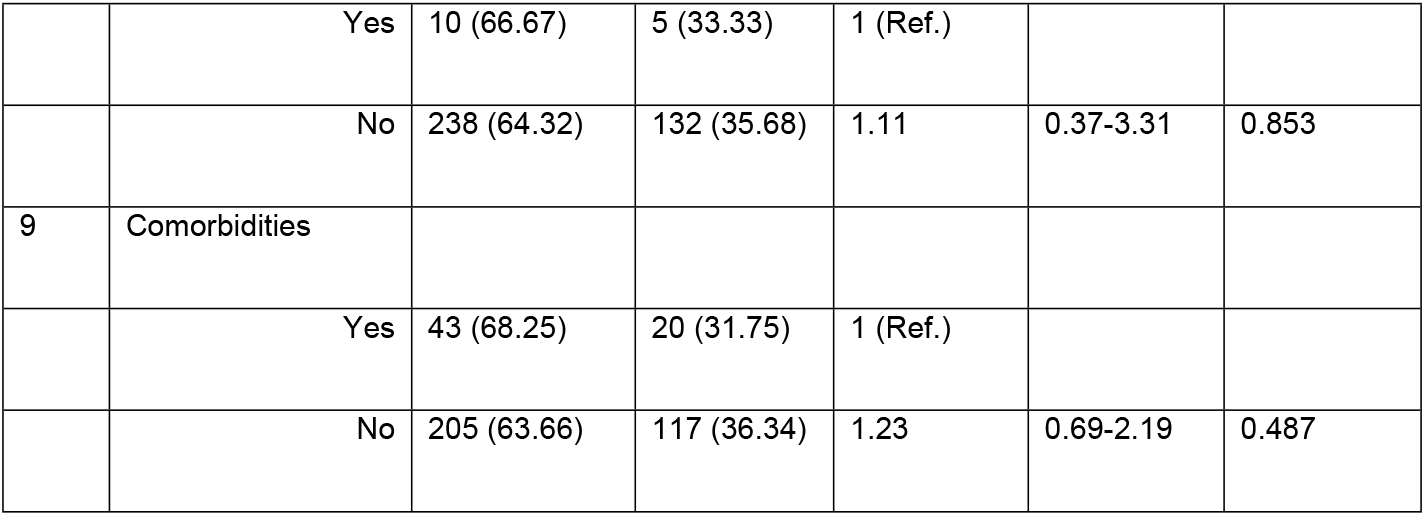
Factors affecting the knowledge of the participants about the rational use of antibiotics.

| SN | Characteristics | Knowledge |  | Binary logistic regression |  |  |
| --- | --- | --- | --- | --- | --- | --- |
|  |  | Moderate to Poor (%) | Good (%) | OR | 95% CI | P-value |
| 1 | Age |  |  |  |  |  |
|  | <40 years | 231(64.17) | 129 (35.83) | 1 (Ref.) |  |  |
| | ≥40 years | 17 (68.00) | 8 (32.00) | 0.84 | 0.35-2.01 | 0.699 |
| 2 | Gender |  |  |  |  |  |
|  | Male | 95 (59.75) | 64 (40.25) | 1 (Ref.) |  |  |
|  | Female | 153 (67.70) | 73 (32.30) | 0.71 | 0.46-1.08 | 0.109 |
| 3 | Education |  |  |  |  |  |
|  | Up to Bachelor | 124 (63.59) | 71 (36.41) | 1 (Ref.) |  |  |
|  | Master & above | 124 (65.26) | 66 (34.74) | 0.93 | 0.61-1.41 | 0.732 |
| 4 | Residency/Area |  |  |  |  |  |
|  | Urban | 200 (65.57) | 105 (34.43) | 1 (Ref.) |  |  |
|  | Rural | 48 (60.00) | 32 (40.00) | 1.27 | 0.77-2.11 | 0.354 |
| 5 | Occupation |  |  |  |  |  |
|  | Not employed/Student | 145 (65.02) | 78 (34.98) | 1 (Ref.) |  |  |
|  | Employed | 103 (63.58) | 59 (36.42) | 1.07 | 0.70-1.62 | 0.770 |
| 6 | Nature of work |  |  |  |  |  |
|  | Health professionals | 143 (59.58) | 97 (40.42) | 1 (Ref.) |  |  |
|  | Others | 105 (72.41) | 40 (27.59) | 0.56 | 0.36-0.88 | 0.011 |
| 7 | Income |  |  |  |  |  |
|  | <50000 | 213 (65.94) | 110 (34.05) | 1 (Ref.) |  |  |
|  | ≥50000 | 35 (56.45) | 27 (43.55) | 1.49 | 0.86-2.60 | 0.153 |
| 8 | Smoking status |  |  |  |  |  |
|  | Yes | 10 (66.67) | 5 (33.33) | 1 (Ref.) |  |  |
|  | No | 238 (64.32) | 132 (35.68) | 1.11 | 0.37-3.31 | 0.853 |
| 9 | Comorbidities |  |  |  |  |  |
|  | Yes | 43 (68.25) | 20 (31.75) | 1 (Ref.) |  |  |
|  | No | 205 (63.66) | 117 (36.34) | 1.23 | 0.69-2.19 | 0.487 |

### Behaviour assessment

The responses are reported in **Supplementary Table 2**. The median behaviour score was 21 (25^th^ to 75^th^ percentiles: 18-23). More than three-fifths of the participants (248, 64.42%) had good behaviour regarding the RUM. The participants with an education master’s degree and/or above had more than two times higher odds of good behaviour (OR 2.55, 95% CI 1.68-3.87). Employed participants had more than two times higher odds of good behaviour (OR 2.90, 95% CI 1.84-4.57) than non-employed or students. The health professionals had 2.38 times higher odds of good behaviour than other professionals (OR 0.42, 95% CI 0.27-0.64). Similarly, participants with an income of more than 50000 Nepali rupees (NRS) had 3.37 times higher odds of good behaviour (OR 3.37, 95% CI 1.65-6.87) (Tables 2 and 4).

**Table 4:**
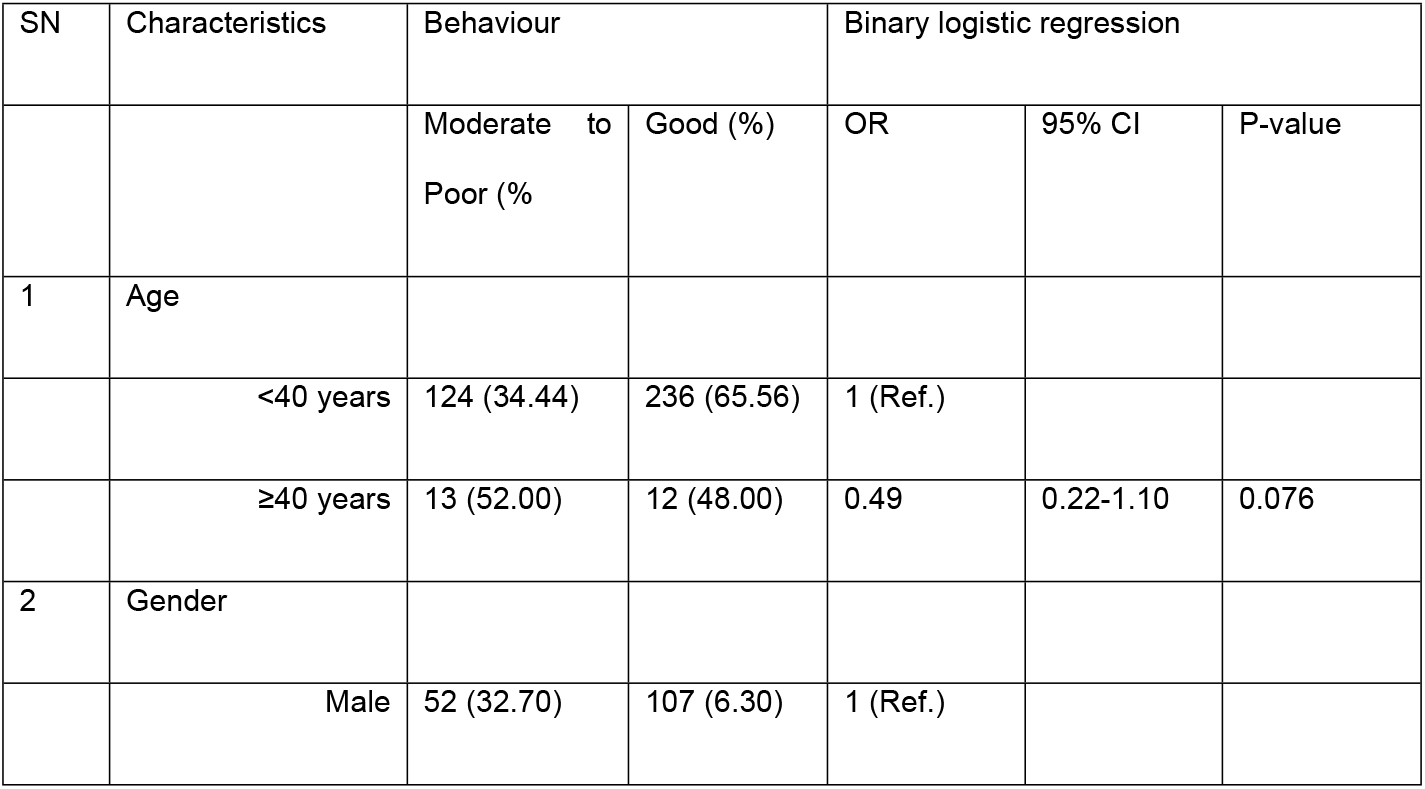
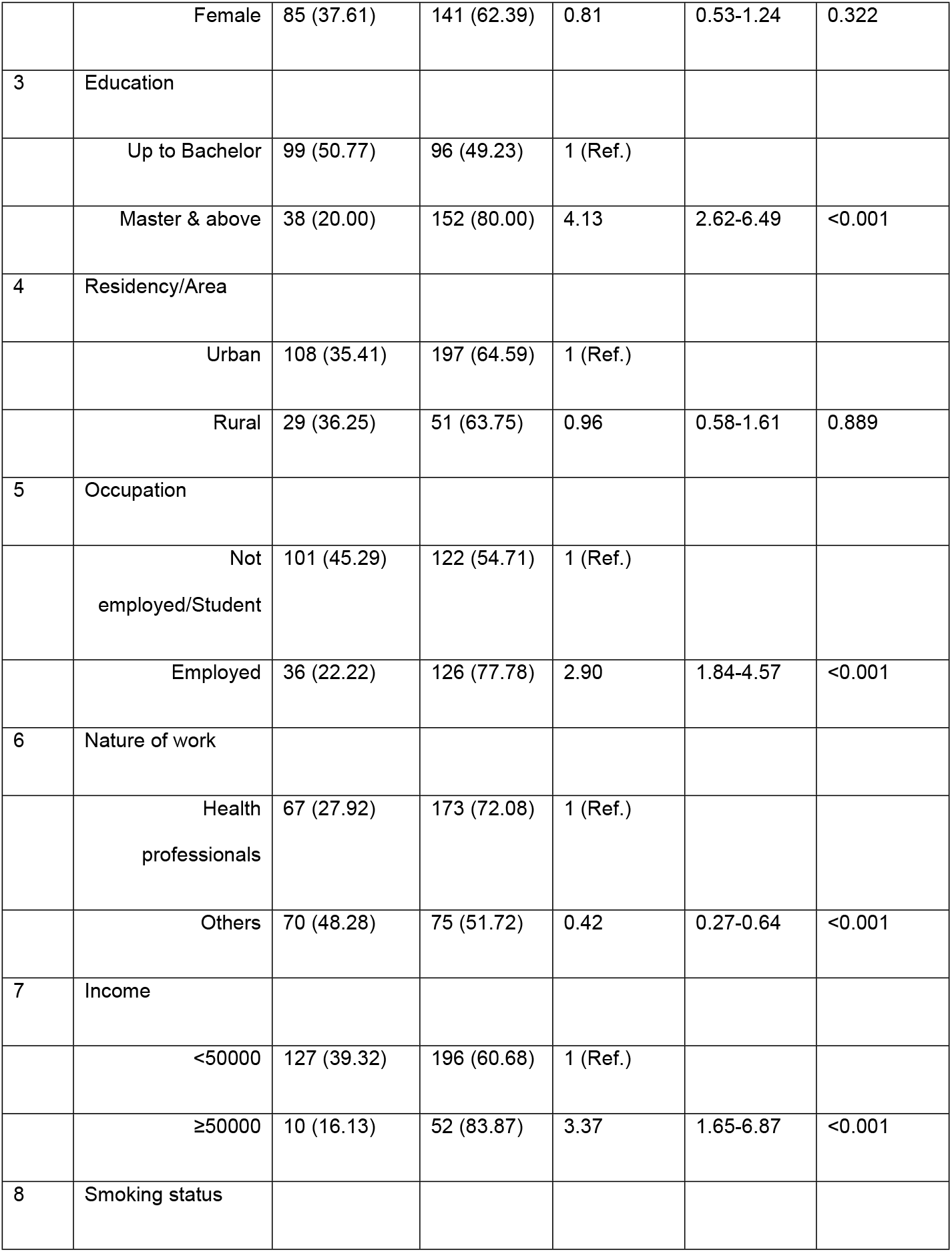
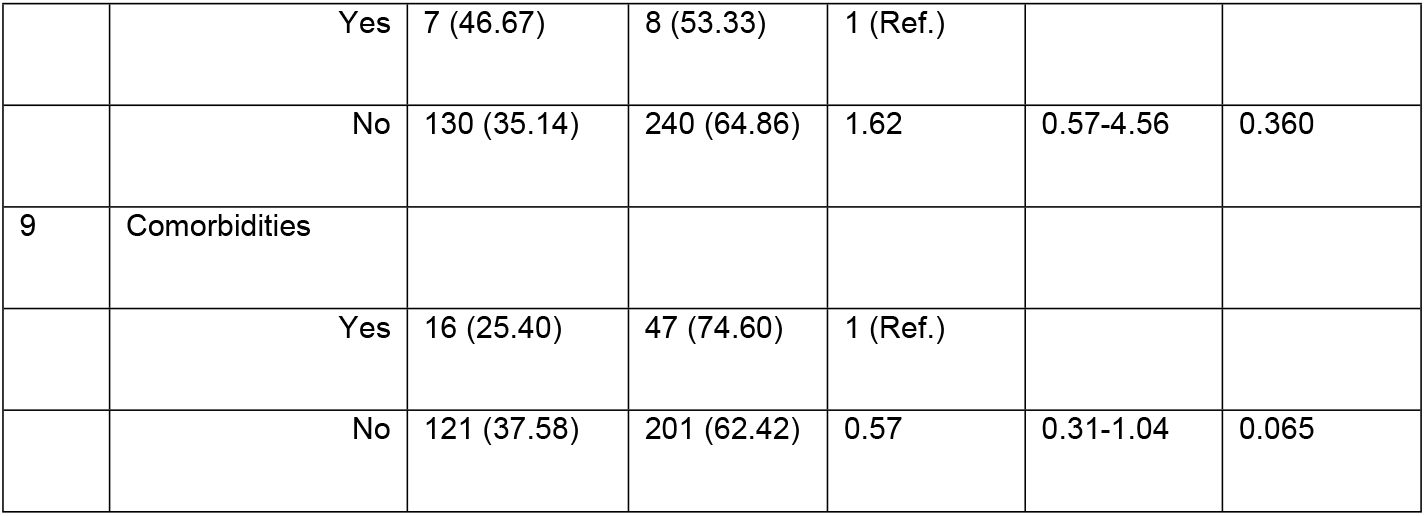
Factors affecting the Behaviour of the participants about the rational use of antibiotics.

| SN | Characteristics | Behaviour |  | Binary logistic regression |  |  |
| --- | --- | --- | --- | --- | --- | --- |
|  |  | Moderate to Poor (%) | Good (%) | OR | 95% CI | P-value |
| 1 | Age |  |  |  |  |  |
|  | <40 years | 124 (34.44) | 236 (65.56) | 1 (Ref.) |  |  |
|  | ≥40 years | 13 (52.00) | 12 (48.00) | 0.49 | 0.22-1.10 | 0.076 |
| 2 | Gender |  |  |  |  |  |
|  | Male | 52 (32.70) | 107 (6.30) | 1 (Ref.) |  |  |
|  | Female | 85 (37.61) | 141 (62.39) | 0.81 | 0.53-1.24 | 0.322 |
| 3 | Education |  |  |  |  |  |
|  | Up to Bachelor | 99 (50.77) | 96 (49.23) | 1 (Ref.) |  |  |
|  | Master & above | 38 (20.00) | 152 (80.00) | 4.13 | 2.62-6.49 | <0.001 |
| 4 | Residency/Area |  |  |  |  |  |
|  | Urban | 108 (35.41) | 197 (64.59) | 1 (Ref.) |  |  |
|  | Rural | 29 (36.25) | 51 (63.75) | 0.96 | 0.58-1.61 | 0.889 |
| 5 | Occupation |  |  |  |  |  |
|  | Not employed/Student | 101 (45.29) | 122 (54.71) | 1 (Ref.) |  |  |
|  | Employed | 36 (22.22) | 126 (77.78) | 2.90 | 1.84-4.57 | <0.001 |
| 6 | Nature of work |  |  |  |  |  |
|  | Health professionals | 67 (27.92) | 173 (72.08) | 1 (Ref.) |  |  |
|  | Others | 70 (48.28) | 75 (51.72) | 0.42 | 0.27-0.64 | <0.001 |
| 7 | Income |  |  |  |  |  |
|  | <50000 | 127 (39.32) | 196 (60.68) | 1 (Ref.) |  |  |
|  | ≥50000 | 10 (16.13) | 52 (83.87) | 3.37 | 1.65-6.87 | <0.001 |
| 8 | Smoking status |  |  |  |  |  |
|  | Yes | 7 (46.67) | 8 (53.33) | 1 (Ref.) |  |  |
|  | No | 130 (35.14) | 240 (64.86) | 1.62 | 0.57-4.56 | 0.360 |
| 9 | Comorbidities |  |  |  |  |  |
|  | Yes | 16 (25.40) | 47 (74.60) | 1 (Ref.) |  |  |
|  | No | 121 (37.58) | 201 (62.42) | 0.57 | 0.31-1.04 | 0.065 |

### Practice assessment

The responses are reported in **Supplementary Table 3**. The median practice score was 38 (25^th^ to 75^th^ percentiles: 34-43). More than two-fifth of the participants (161,41.82%) demonstrated a good practice regarding the RUM. The participants with an education master’s degree and/or above had more than four times higher odds of good practice (OR 2.55, 95% CI 1.68-3.87). Employed participants had more than two times higher odds of good practice (OR 2.56, 95% CI 1.68-3.88) than non-employed or students. Similarly, participants with an income of more than 50000 NRS had 2.58 times higher odds of good practice (OR 2.58, 95% CI 1.47-4.50) (Tables 2 and 5).

**Table 5:**
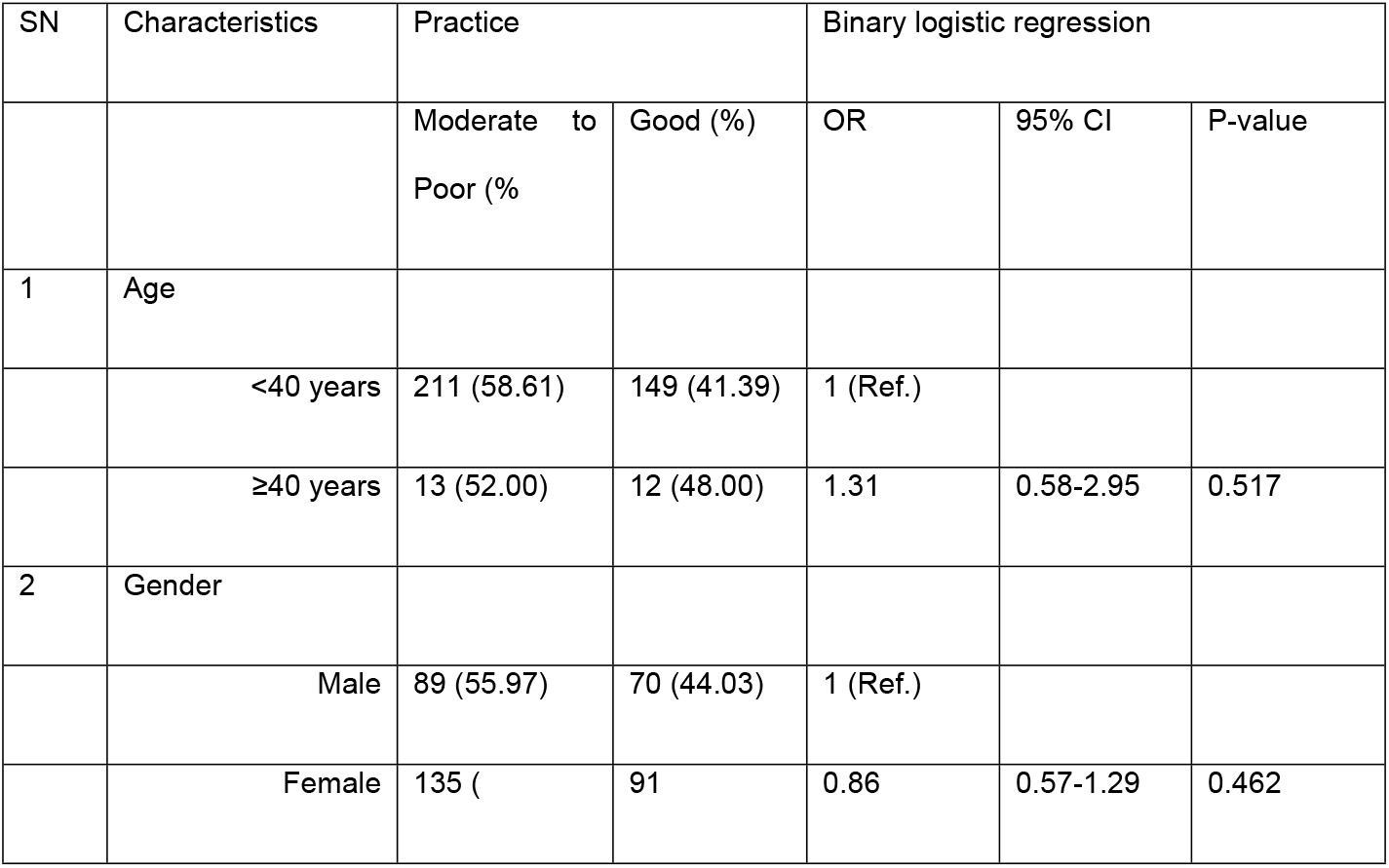
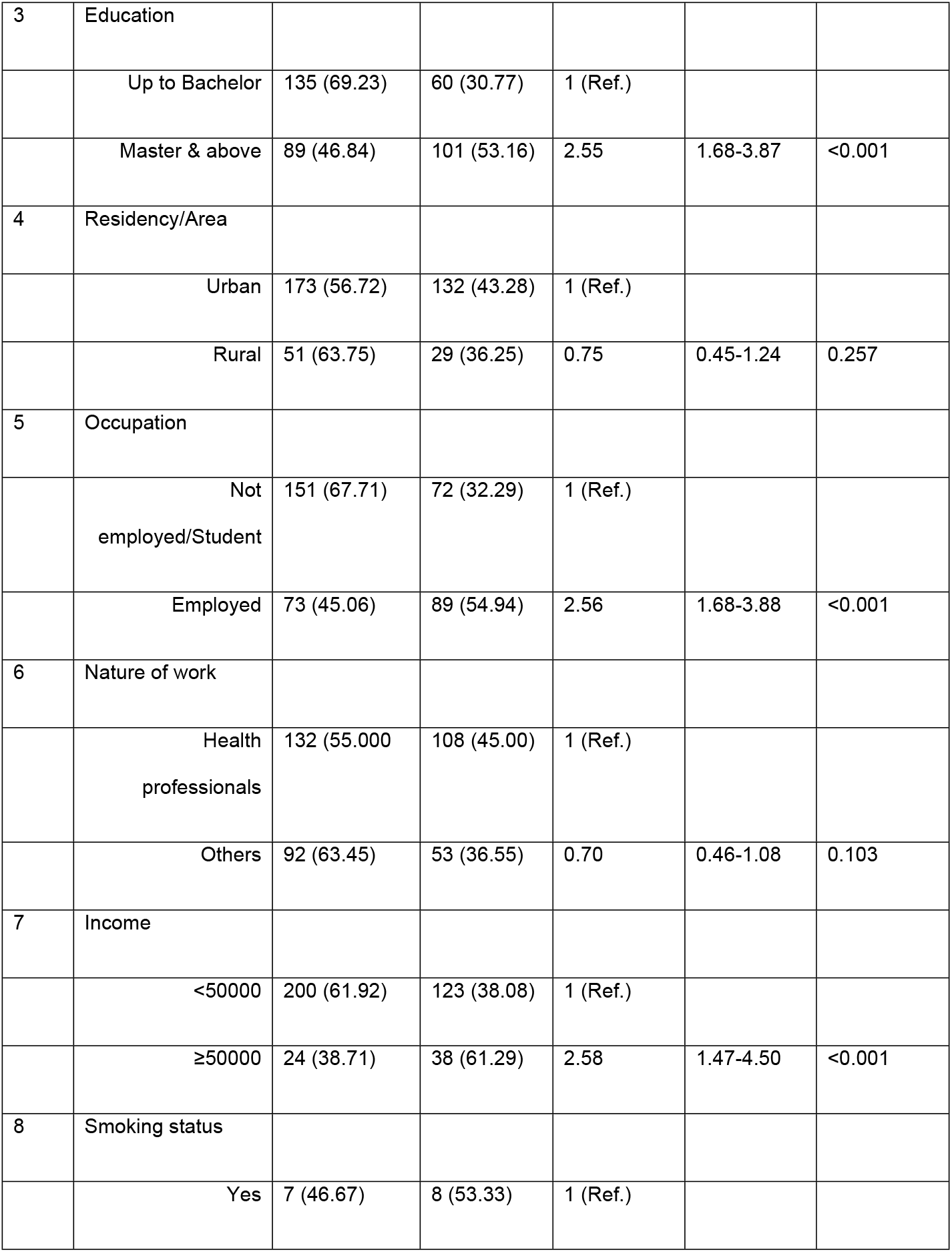
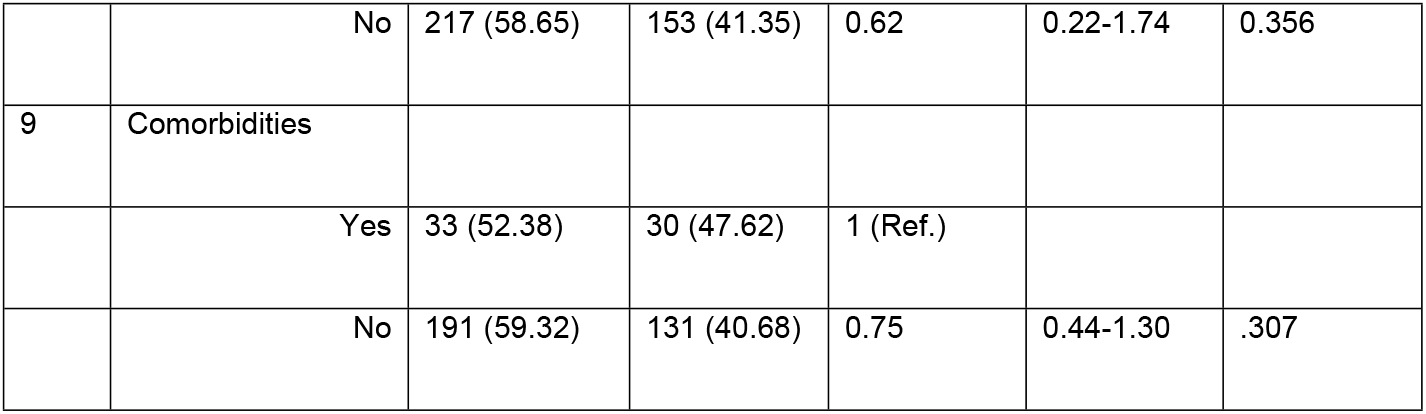
Factors affecting the practice of the participants regarding the rational use of antibiotics.

### Distribution of knowledge, behaviour, and practice scores based on the education level

Those participants with master’s degree and above educational levels had higher median behaviour and practice scores as compared to those with lower educational levels. However, there was no statistical difference across both groups in terms of knowledge, behaviour, and practice scores (Figure 2).

**Figure 2:**
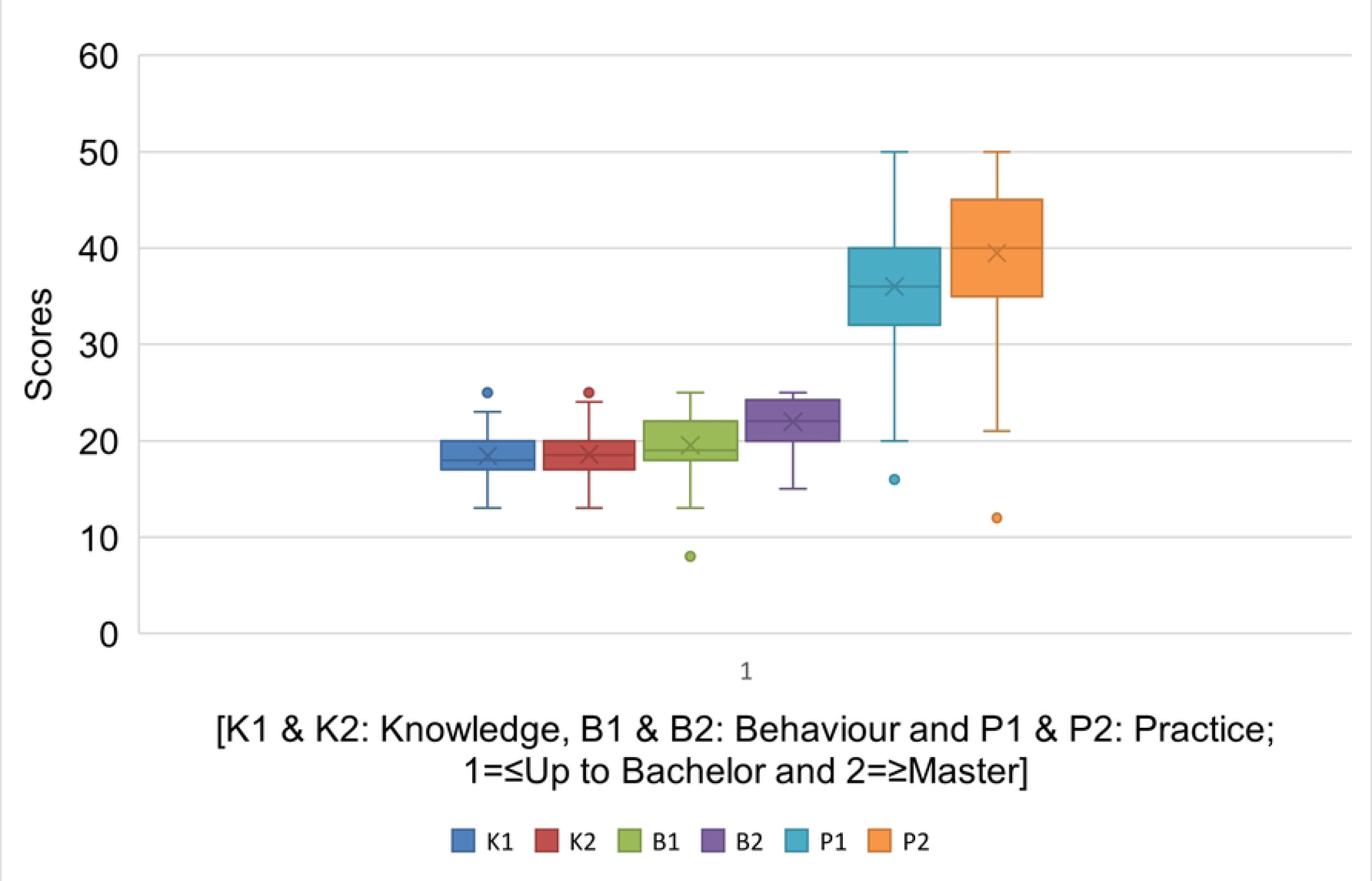
Box plots of the distribution of knowledge, behaviour, and practice scores based on education level.

### Distribution of knowledge, behaviour, and practice scores based on areas of work

There was no statistical difference across both groups (health professionals versus others) in terms of knowledge, behaviour, and practice scores (Figure 3).

**Figure 3:**
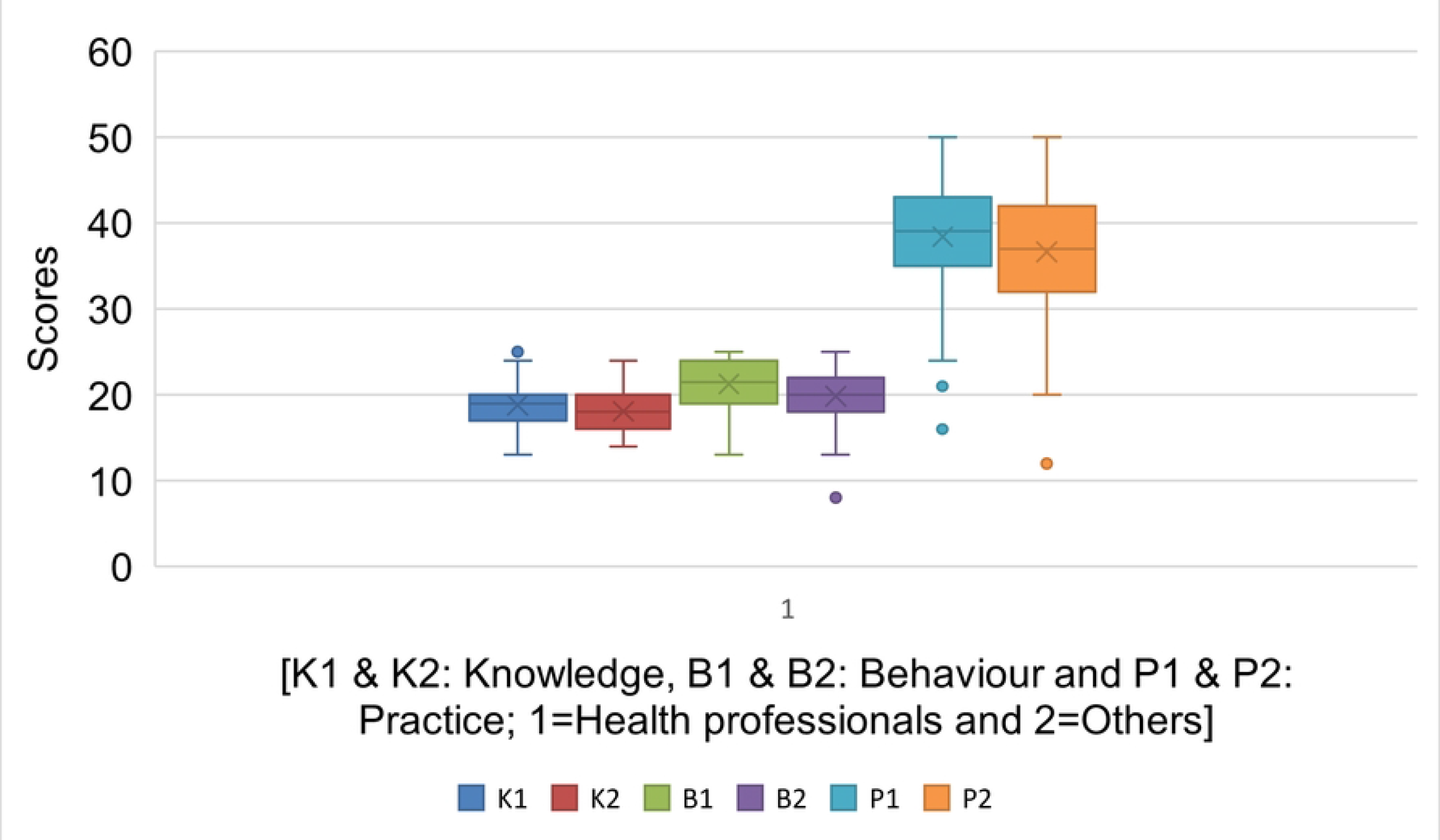
Box plots of the distribution of knowledge, behaviour, and practice scores based on the area of work (health professionals’ v/s others)

### Correlation between scores

There was significant correlation between knowledge and behaviour scores (Spearman’s rho: 0.331; P-value = <0.001, 95% CI=0.24-0.42). Similarly, the knowledge and practice scores (Spearman’s rho: 0.259; P-value <0.001, 95% CI=0.16-0.35), and behaviour and practice scores were positively correlated (Spearman’s rho: 0.618; P-value = <0.001, 95% CI=0.55-0.68).

## Discussion

Accumulative risk due to AMR caused by irrational use of antibiotics is a global health concern. Such risks lead to health and economic ramifications, including preventable deaths, increased healthcare expenditure, and higher levels of healthcare overheads [17].”Targeted spectrum” antibiotic use in appropriate dose and duration can prevent the evolving AMR [18].

The factors such as the lack of surgical prophylaxis protocols, documentation of AMR, and associated mortality indicated the need for investment in protocol development and robust surveillance in a similar national-level study conducted among various groups of the population [19]. Our study helped us understand the difference in knowledge, behaviour, and practice among the population of different socioeconomic backgrounds.

A study performed in Saudi Arabia among general population showed insufficient knowledge regarding antibiotic safety and unethical medical use of antibiotic [20]. However, some studies done in China and UAE demonstrated better knowledge, attitude, and practice of antibiotic use in a specific group of people like medical students than the general population, which is in line with the findings of our study [21,22].

Obtaining antibiotics without prescription is a common practice all over the world. The overall prevalence of antibiotic self-medication in developing countries was found to be 38.8 % in a metanalysis performed by Ocan et al. [23]. The overall inappropriate antibiotics use in a study performed in Northwest Ethiopia was 30.9% of which 18% was used for self-medication and 12.9% was used for treatment of family member [24]. A study done in LMICs like Eritrea showed the extent of dispensing antibiotic from the retail outlets without prescription to 87% [25]. In our study, about one third participants believed that antibiotics can be obtained from a pharmacy without a doctor’s prescription like other over-the-counter (OTC) drugs. Further, 15.58% took antibiotics without prescription. Self-use of antibiotics was found 15.5% in undergraduate students of Nigeria similar to our study, 30.2% among the metropolitan people of Thailand, and 23.7% in South India [26-28].

In our study, 26 % stopped taking antibiotics as soon as the symptoms subsided. It is similar to the study in Northwest Ethiopia where 27% of the participants discontinued their antibiotics once the symptoms subsided [29]. This percentage is higher (67%) in Saudi Arabia [20].

A considerable number of patients approach directly to pharmacy/pharmacists for resolving their medical issues and they exert great pressure on such professionals to obtain antibiotics without a prescription [30]. Thus, pharmacists in community can play a role as first-line health professionals in effective patient counseling and public awareness regarding use of prescription antibiotics and adverse effects of antibiotic misuse.

In our study, it was also found that about 15.07% requested the physicians to prescribe medications without culture report. In a study done in Pakistan, 38.1% had negative attitude to the statement that there is a need for culture analysis before dispensing antibiotics [31]. Although policy guidelines demand the use of antibiotics based on the identification of causative agents, the empirical and OTC use of antibiotics are high in practice [32]. Performing culture is crucial to understand the resistance pattern in the patient. Knowing the trends in sensitivity and resistance patterns by conducting culture and sensitivity tests can assist physicians and policymakers in making better judgments about how to address potential resistance [33,34].

Knowledge regarding the use of antibiotics against the minor ailments like common cold, flu, and cough seemingly found poor as 32.21% of the population thought of using antibiotics for the ailments while 27.53% actually took them which is in line with the results of a study done in Kuwait, Jordan and UK [35-37]. This is probably due to inadequate knowledge of the dosage and duration of the antibiotic use. Patients who do not complete treatment are more likely to relapse, develop resistance, and need another treatment (13). Access to antibiotics is a major concern in many countries worldwide especially LMICs [38]. The findings of our study concurred with the statement as about 45.20% of respondents mentioned that they have taken antibiotics of a similar group because the prescribed one was not available. The reasons of inaccessibility is attributed to the very low economic status of a large number of nations, inappropriate use, high cost of the most recent and efficacious antibiotics, extensive OTC usage, an increasing number of counterfeit drugs, and a dramatic increase in AMR [39].

Of the participants, 31.95 % took low doses of antibiotics due to fear of side effects in this study and about one-fourth (18.96%) took the leftover antibiotics contrary to which a study done in Germany presented that 88.7% received advice from a doctor or pharmacist on how to take the prescribed antibiotics [40]. However, the use of leftover drugs is widespread mainly in LMICs where many types of drugs are sold without prescriptions [28]. It is the responsibility of medical professionals to provide proper counseling on usage such as dose, frequency of dose, treatment course, and the harmful effects of misuse [41]. However, it is lacking in LMICs like Nepal, owing to the doctor’s less time contribution to the patient as a result of the low doctor-to-patient ratio. Rampant antibiotic use and ignorance of people about the complete knowledge of the course of antibiotics, their side effects, standard acceptable dosage limits, and antibiotic overdose issues are the potential reasons for inappropriate or incorrect treatment which can lead to AMR issues and increased morbidity [42]. The lack of implementation of uniform and nationwide guideline on how patients should safely dispose of their leftover medications can result in people overusing them, and giving them away to family, friends, or charity centers [43]. One of the easiest ways to reduce the use of leftovers could be the shortening course of antibiotics to 3 or 5 days.

Our study findings showed a significant association between the variables, nature of work (compared to health professionals and others), and knowledge and behaviour regarding antibiotic use among respondents which corresponded with the study done in Nepal and Hongkong [44,45]. Our study showed that the knowledge and practice of antibiotics use and AMR is significantly dependent on the education level of the respondents which align with the findings of a study done in Lithuania where people with an education level of a college degree and above had better knowledge, more appropriate behaviours, and better practices [46]. Similarly, the level of income was significantly and synonymously associated with the behaviour and practice of antibiotic use in our study which contrasts with a study performed recently but a similar finding was seen in one of the studies in Saudi Arabia [47,48]. The same study demonstrated a significant association between the nature of work and behaviour and practice on antibiotic use like our study where health professionals were better informed about the correct use of antibiotics. The remaining demographic variables did not significantly influence the knowledge, behaviour, and practice of antibiotic use. The exact reason for this is currently not clear, but it can be postulated that the different geographic locations, healthcare regulations, and policies of the nation may be a contributing factor, although future research is needed to investigate.

Important concern for antibiotic resistance is the irrational use of antibiotics for plants and food animals. Antibiotic use in various industries; for instance, in the United States, agriculture, farming, and aquaculture account for around 80% of antibiotic usage [49]. In LMICs, antibiotics are frequently employed against rice to combat mites and insects that are not affected by those antibiotics [50]. The potential hazard to human health arising from incorrect antibiotic use in food animals is also high [51]. Hence, the agricultural use of antibiotics should also be acknowledged as one of the key factors in the emergence of resistant microorganisms.

AMR could be addressed by adopting One Health (OH) approach that brings together humans, animals, and the environmental sector. The government of Nepal has also adopted the approach as highlighted by GAP [10,52]. However, there are challenges such as a lack of separate institutional setup, lack of awareness among professionals in human and animal health, and environment sectors, unclear job and responsibilities, and regulatory mechanisms [53]. Training and seminars from the policy-making level is highly recommended. Public dissemination of information using television, newspapers, and social media is an effective method of increasing health literacy.

Our findings can be considered in the context of several limitations. First, the results contain self-reported, online reports which may not reflect the actual behaviour. Second, the response to the survey was based on the author’s network, and as such, they might ignore valuable comments from other people who had not been surveyed. Third, most of the study participants had a high academic level. Fourth, most participants belonged to urban areas with better access to information on antibiotics. However, our findings can provide helpful information for determining antibiotic use awareness.

## Conclusion

Antibiotics misuse is highly prevalent in the low-and middle-income countries. Lack of execution of existing laws and unawareness of the public is responsible for the rampant use of antibiotics. Our study reflected on various aspects of people’s knowledge, attitude, and practice regarding antibiotics use in low and middle-income countries. The findings of this study may serve as a baseline data to understand people’s perception regarding AMR in LMICs. Our study found a significant association between the nature of work; health professionals versus others; and knowledge, behaviour, and practice of antibiotic use. In addition, a significant association was found between the level of education and income and the behaviour and practice regarding antibiotics use. This study demands effective legislature - strict enforcement of the drug act and proper implementation of other relevant plans and policies. Studies focusing on the effective public education and awareness are highly recommended. Further studies are also necessary to evaluate the effects of awareness programs on antibiotics use that hep contain antibiotics and thus antimicrobial resistance.

## Data Availability

All relevant data are within the manuscript and its Supporting Information files.

## Notes

### Competing Interest Statement

The authors have declared no competing interest.

### Funding Statement

The author(s) received no specific funding for this work.

## References

1. Alhomoud F, Almahasnah R, Alhomoud FK. “You could lose when you misuse” -factors affecting over-the-counter sale of antibiotics in community pharmacies in Saudi Arabia: a qualitative study. BMC Health Serv Res. Dec 3 2018;18(1):915.

2. Auta A, Hadi MA, Oga E, et al. Global access to antibiotics without prescription in community pharmacies: A systematic review and meta-analysis. Journal of Infection. 2019;78(1):8–18.

3. World Health Organization. Global Action Plan on Antimicrobial Resistance. Available at: https://www.who.int/antimicrobial-resistance/global-action-plan/en/. Accessed 11 July 2020.

4. World Health Organization. WHO global strategy for containment of antimicrobial resistance: World Health Organization; 2001

5. Dahal RH, Chaudhary DK. Microbial Infections and Antimicrobial Resistance in Nepal: Current Trends and Recommendations. Open Microbiol J. 2018;12:230– 42.

6. Tenover FC. Mechanisms of antimicrobial resistance in bacteria. The American journal of medicine. 2006;119(6):S3–S10.

7. Holmes AH, Moore LS, Sundsfjord A, et al. Understanding the mechanisms and drivers of antimicrobial resistance. The Lancet 2016;387(10014):176–187.

8. Boucher HW, Talbot GH, Bradley JS, et al. Bad bugs, no drugs: no ESKAPE! An update from the Infectious Diseases Society of America. Clinical infectious diseases. 2009;48(1):1–12.

9. Prestinaci F, Pezzotti P, Pantosti A. Antimicrobial resistance: a global multifaceted phenomenon. Pathogens global health 2015;109(7):309–318.

10. Khadka, S., and S. Giri. “ Addressing Antimicrobial Resistance in Nepal: A Call for Collaborative Efforts”. Europasian Journal of Medical Sciences, Vol. 3, no. 2, July 2021, pp. 1–3, doi:10.46405/ejms.v3i2.354.

11. Malik B, Bhattacharyya S. Antibiotic drug-resistance as a complex system driven by socio-economic growth and antibiotic misuse. Sci Rep. 2019 Jul 5;9(1):9788.

12. Ventola CL. The antibiotic resistance crisis: part 1: causes and threats. P T. 2015 Apr;40(4):277–83.

13. World Health Organization. Antimicrobial resistance global report on surveillance: 2014 summary: World Health Organization; 2014.

14. MoHP. National Antimicrobial ResistanceContainment Action Plan Nepal. MoHP. 2016.https://www.flemingfund.org/wp-content/

15. Jan P. Vandenbroucke, Erik von Elm, Douglas G. Altman, Peter C. Gøtzsche, Cynthia D. Mulrow, Stuart J. Pocock et.al, Strengthening the Reporting of Observational Studies in Epidemiology (STROBE): Explanation and elaboration,International Journal of Surgery, Volume 12, Issue 12, 2014, Pages 1500–1524, ISSN 1743-9191, https://doi.org/10.1016/j.ijsu.2014.07.014.

16. Seid, M.A., Hussen, M.S. Knowledge and attitude towards antimicrobial resistance among final year undergraduate paramedical students at University of Gondar, Ethiopia. BMC Infect Dis 18, 312 (2018). https://doi.org/10.1186/s12879-018-3199-1

17. Byrne MK, Miellet S, McGlinn A, Fish J, Meedya S, Reynolds N, et al. The drivers of antibiotic use and misuse: the development and investigation of a theory driven community measure. BMC Public Health. 2019 Oct 30;19(1):1425.

18. Lieberman JM. Appropriate antibiotic use and why it is important: the challenges of bacterial resistance: The Pediatric Infectious Disease Journal. 2003 Dec;22(12):1143–51.

19. Rijal, K.R., Banjara, M.R., Dhungel, B. et al. Use of antimicrobials and antimicrobial resistance in Nepal: a nationwide survey. Sci Rep 11, 11554 (2021). https://doi.org/10.1038/s41598-021-90812-4

20. Al-Shibani N, Hamed A, Labban N, Al-Kattan R, Al-Otaibi H, Alfadda S. Knowledge, attitude and practice of antibiotic use and misuse among adults in Riyadh, Saudi Arabia. Saudi Med J. 2017 Oct;38(10):1038–44.

21. Huang Y, Gu J, Zhang M, Ren Z, Yang W, Chen Y, et al. Knowledge, attitude and practice of antibiotics: a questionnaire study among 2500 Chinese students. BMC Med Educ. 2013 Dec 9;13(1):163.

22. Jairoun A, Hassan N, Ali A, Jairoun O, Shahwan M. Knowledge, attitude and practice of antibiotic use among university students: a cross sectional study in UAE. BMC Public Health. 2019 May 6;19(1):518.

23. Ocan M, Obuku EA, Bwanga F, Akena D, Richard S, Ogwal-Okeng J, et al. Household antimicrobial self-medication: a systematic review and meta-analysis of the burden, risk factors and outcomes in developing countries. BMC Public Health. 2015 Aug 1;15(1):742

24. Gebeyehu E, Bantie L, Azage M. Inappropriate Use of Antibiotics and Its Associated Factors among Urban and Rural Communities of Bahir Dar City Administration, Northwest Ethiopia. PLOS ONE. 2015 Sep 17;10(9):e0138179

25. Russom M, Bahta M, Debesai M, Bahta I, Kessete A, Afendi A, et al. Knowledge, attitude and practice of antibiotics and their determinants in Eritrea: an urban population-based survey. BMJ Open. 2021 Sep 1;11(9):e046432.

26. Akande-Sholabi W, Ajamu AmenT, Adisa R. Prevalence, knowledge and perception of self-medication practice among undergraduate healthcare students. Journal of Pharmaceutical Policy and Practice. 2021 Jun 10;14(1):49.

27. Chautrakarn S, Khumros W, Phutrakool P. Self-Medication With Over-the-counter Medicines Among the Working Age Population in Metropolitan Areas of Thailand. Frontiers in Pharmacology [Internet]. 2021 [cited 2022 Jun 18];12. Available from: https://www.frontiersin.org/article/10.3389/fphar.2021.726643

28. Kumar V, Mangal A, Yadav G, Raut D, Singh S. Prevalence and pattern of self-medication practices in an urban area of Delhi, India. Medical Journal of Dr DY Patil University. 2015 Jan 1;8(1):16.

29. Gebeyehu E, Bantie L, Azage M. Inappropriate Use of Antibiotics and Its Associated Factors among Urban and Rural Communities of Bahir Dar City Administration, Northwest Ethiopia. PLOS ONE. 2015 Sep 17;10(9):e0138179.

30. Llor C, Monnet D, Cots J. Small pharmacies are more likely to dispense antibiotics without a medical prescription than large pharmacies in Catalonia, Spain. Euro Surveill. 2010 Aug 12;15(32):19635.

31. Hashmi F, Khadka S, Khan M, Khan S, Saeed H, Islam M, et al. Antimicrobial Dispensing Sans Legitimate Prescription from Community Pharmacies in Lahore, Pakistan: Implications for Antimicrobial Resistance. 2021

32. Pokharel S, Raut S, AdhikariBTackling antimicrobial resistance in low-income and middle-income countries BMJ Global Health 2019;4:e002104

33. Gopalakrishnan R, Sureshkumar D. Changing trends in antimicrobial susceptibility and hospital acquired infections over an 8 year period in a tertiary care hospital in relation to introduction of an infection control programme. The Journal of the Association of Physicians of India. 2010;

34. Jorak A, Keihanian F, Saeidinia A, Heidarzadeh A, Saeidinia F. A Cross Sectional Study on Knowledge, Attitude and Practice of Medical Students Toward Antibiotic Resistance and its Prescription, Iran. Advances in Environmental Biology. 2014 Jan 1;8:675–81

35. Awad AI, Aboud EA. Knowledge, Attitude and Practice towards Antibiotic Use among the Public in Kuwait. PLOS ONE. 2015 Feb 12;10(2):e0117910

36. Shehadeh M, Suaifan G, Darwish RM, Wazaify M, Zaru L, Alja’fari S. Knowledge, attitudes and BEHAVIOUR regarding antibiotics use and misuse among adults in the community of Jordan. A pilot study. Saudi Pharmaceutical Journal. 2012 Apr;20(2):125–33.

37. Cliodna A. M. McNulty, Paul Boyle, Tom Nichols, Peter Clappison, Peter Davey, The public’s attitudes to and compliance with antibiotics, Journal of Antimicrobial Chemotherapy, Volume 60, Issue suppl_1, August 2007, Pages i63–i68, https://doi.org/10.1093/jac/dkm161

38. Bush K, Courvalin P, Dantas G, Davies J, Eisenstein B, Huovinen P, et al. Tackling antibiotic resistance. Nat Rev Microbiol. 2011 Nov 2;9(12):894–6

39. Carlet J, Pittet D. Access to antibiotics: a safety and equity challenge for the next decade. Antimicrob Resist Infect Control. 2013 Jan 10;2:1

40. Salm F, Ernsting C, Kuhlmey A, Kanzler M, Gastmeier P, et al. (2018) Antibiotic use, knowledge and health literacy among the general population in Berlin, Germany and its surrounding rural areas. PLOS ONE 13(2): e0193336. https://doi.org/10.1371/journal.pone.0193336

41. Rather IA, Kim BC, Bajpai VK, Park YH. Self-medication and antibiotic resistance: Crisis, current challenges, and prevention. Saudi Journal of Biological Sciences. 2017 May 1;24(4):808–12

42. Michael CA, Dominey-Howes D, Labbate M. The Antimicrobial Resistance Crisis: Causes, Consequences, and Management. Frontiers in Public Health [Internet]. 2014 [cited 2022 Mar 1];2. Available from: https://www.frontiersin.org/article/10.3389/fpubh.2014.00145

43. AlAzmi A, AlHamdan H, Abualezz R, Bahadig F, Abonofal N, Osman M. Patients’ Knowledge and Attitude toward the Disposal of Medications. Journal of Pharmaceutics. 2017 Oct 10;2017:e8516741

44. Nepal A, Hendrie D, Robinson S, Selvey LA. Knowledge, attitudes and practices relating to antibiotic use among community members of the Rupandehi District in Nepal. BMC Public Health. 2019 Nov;19(1):1558. DOI: 10.1186/s12889-019-7924-5. PMID: 31771595; PMCID: PMC6880381

45. You JH, Yau B, Choi KC, Chau CT, Huang QR, Lee SS. Public knowledge, attitudes and behaviour on antibiotic use: a telephone survey in Hong Kong. Infection. 2008;36(2):153–157. doi:10.1007/s15010-007-7214-5

46. Pavyde E, Veikutis V, Maciuliene A, Maciulis V, Petrikonis K, Stankevičius E. Public Knowledge, Beliefs and Behaviour on Antibiotic Use and Self-Medication in Lithuania. IJERPH. 2015 Jun 17;12(6):7002–16.

47. Mallah N, Orsini N, Figueiras A, Takkouche B. Income level and antibiotic misuse: a systematic review and dose–response meta-analysis. Eur J Health Econ [Internet]. 2021 Nov 30 [cited 2022 Feb 28]; Available from: https://doi.org/10.1007/s10198-021-01416-8

48. Alnasser AHA, Al-Tawfiq JA, Ahmed HAA, Alqithami SMH, Alhaddad ZMA, Rabiah ASM, et al. Public knowledge, attitude and practice towards antibiotics use and antimicrobial resistance in Saudi Arabia: A web-based cross-sectional survey. J Public Health Res. 2021 Jul 23;10(4):2276

49. Manyi-Loh C, Mamphweli S, Meyer E, Okoh A. Antibiotic Use in Agriculture and Its Consequential Resistance in Environmental Sources: Potential Public Health Implications. Molecules. 2018 Mar 30;23(4):795.

50. Taylor, P., Reeder, R. Antibiotic use on crops in low and middle-income countries based on recommendations made by agricultural advisors. CABI Agric Biosci 1, 1 (2020). https://doi.org/10.1186/s43170-020-00001-y

51. Landers TF, Cohen B, Wittum TE, Larson EL. A review of antibiotic use in food animals: perspective, policy, and potential. Public Health Rep. 2012 Jan-Feb;127(1):4–22. doi: 10.1177/003335491212700103. PMID: 22298919; PMCID: PMC3234384.

52. Acharya K. National Action Plan forAntimicrobial Resistance in Nepal: Possibility ofTranslating Idea into Reality. Open Microbiol J.2020;14:38–9. Global action plan onantimicrobialresistance. WHO. 2015 [cited 2021 Nov 2]. https://www.who.int/antimicrobial-resistanc/global-action-plan/en/

53. Acharya KP, Karki S, Shrestha K, Kaphle K. One health approach in Nepal: Scope,opportunities and challenges. One Heal (Amsterdam, Netherlands). 2019 Dec;8:100101. https://doi.org/10.1016/j.onehlt.2019.100101

